# Primary aldosteronism results in a decline estimated glomerular filtration rate independent of blood pressure: evidence from a case-control and mendelian randomization study

**DOI:** 10.1101/2024.09.08.24313278

**Authors:** Mingjie Xu, Boteng Yan, Mingli Li, Yushuang Wei, Xihui Jin, Xiaoyou Mai, Hui Liang, Haiyun Lan, Wenchao Xie, Tianjiao Pang, Guijian Pang, Qiang Lin, Yifeng Chen, Zeguang Zhou, Yongxian Wu, Xinyang Long, Shengzhu Huang, Chaoyan Tang, Zengnan Mo

## Abstract

**Background:** Primary aldosteronism (PA) is the predominant cause of secondary hypertension, leading to cardiovascular and renal damage via mechanisms such as oxidative stress and fibrosis. However, current epidemiology findings on the association between PA and estimated glomerular filtration rate (eGFR) remain inconsistent.

**Methods:** A 1:1 sex- and age-matched case-control study was conducted among participants with PA, essential hypertension (EH), and normotension, with 204 participants in each group. Multiple linear regression was used to explore the correlations of PA, plasma aldosterone concentration (PAC), plasma renin concentration (PRC), and the aldosterone-to-renin ratio (ARR) with eGFR. Additionally, we performed a bidirectional two-sample mendelian randomization (MR) analysis to assess the causal relationship between PA and eGFR based on public genome wide association study (GWAS) databases, and established a multivariable MR (MVMR) analysis to further explore whether the causal effect of PA on eGFR decline independent of systolic (SBP) or diastolic blood pressure (DBP).

**Results:** Multiple linear regression model showed that PA was associated with a decline eGFR (β = -0.234, [95% CI, -0.099, -0.039], P<0.001) after adjusted potential confounders. When stratified the PA patients into three groups according to the levels of PAC, PRC and ARR, patients in the highest PAC groups (β = -0.146 [95% CI, -0.093, -0.008], P =0.021), the lowest PRC group (β = -0.127 [95% CI, -0.084, -0.004], P =0.033), and the highest ARR group (β = -0.147 [95% CI, -0.092, -0.009], P =0.017) had much lower eGFR compared to the EH group. The inverse associations mentioned above remained significant even further adjusted for SBP or DBP, respectively. Besides, MR results indicated that genetically predicted PA was causally associated with a decline eGFR (β =-6.671×10^-4^[95% CI,-1.291×10^-4^,-4.328×10^-5^], P = 0.036), consistent effects were further detected in SBP (β =-1.121×10^-3^ [95% CI,-2.132×10^-3^,-1.110×10^-4^], P = 0.029) or DBP (β =-1.542×10^-3^,[95% CI,-2.693×10^-3^,-3.912×10^-4^], P = 0.008) adjusted model using MVMR analysis.

**Conclusion:** Our study indicates that PA is causally associated with lower eGFR independent of blood pressure, and the adverse effects might be greater than negative controls or EH patients.

## Introduction

Primary aldosteronism (PA) is the leading cause of secondary hypertension, detected in 5%-13% of the hypertensive population ^[1]^. Excess aldosterone leads to the overactivation of the mineralocorticoid receptor, subsequently resulting in target organ damage through mechanisms such as fibrosis, oxidative stress, and tissue remodeling ^[2-4]^. Renal dysfunction is a principal complication in PA patients, where excessive mineralocorticoid receptor activation induces renal inflammation, mesangial cell proliferation, interstitial fibrosis, and glomerulosclerosis^[5,6]^, ultimately impairing renal function. Compared to patients with essential hypertension (EH), those with PA exhibit a higher incidence of chronic kidney disease and face a significantly increased risk of disease progression^[7,8]^.

The estimated glomerular filtration rate (eGFR) is universally recognized as the optimal indicator for assessing renal function and is a crucial prognostic factor for the progression to end-stage renal disease^[9,10]^, It is also strongly associated with clinical outcomes, including cardiovascular events, the initiation of dialysis, and all-cause mortality^[11,12]^. However, the correlation between PA and eGFR is still ambiguous, with prior limited studies presenting conflicting conclusions. A case-control study in 2009 that compared PA with EH discovered that patients with PA had higher serum creatinine levels and lower GFR than those with EH, and identified plasma aldosterone concentration (PAC) as an independent risk factor for decreased GFR^[7]^. Ribstein et al. reported no difference in baseline GFR between PA and EH patients^[13]^, whereas other studies observed that PA patients exhibited higher baseline eGFR levels than EH patients during long-term follow-up^[14,15]^. These contradictory results underscore the need for further research to clarify the precise relationship between PA and eGFR, especially across different geographic and demographic contexts. Mendelian randomization (MR) utilizes genetic variants associated with modifiable exposures (or risk factors) to evaluate their potential causal relationships with outcomes^[16]^. Since genetic variants are randomly assigned at conception, this method effectively eliminates potential sources of error and establishes a clear causal chain. Consequently, MR minimizes potential biases arising from confounding factors and reverse causation, thereby enhancing the reliability of the results^[17]^.However, current studies primarily focus on correlations with clinical physiological markers, with few exploring the causal relationship between PA and eGFR from a genetic perspective.

We therefore conducted a cross-sectional survey in the southwestern region of China, measuring PAC, plasma renin concentration (PRC), the aldosterone-to-renin ratio (ARR), and renal function of participants, while also collecting their medical histories and other relevant clinical data. A case-control study was designed, including groups of patients with PA, EH, and negative control (NC). By analyzing these three groups, we could more accurately assess the impact of PA on renal function. Utilizing genome wide association study (GWAS) public databases, we employed a two-sample Mendelian randomization approach to explore the potential causal relationship between PA and eGFR.

## Materials and methods

### Study design and participants

This cross-sectional study recruited participants from September 2022 to March 2024 at the First Affiliated Hospital of Guangxi Medical University and the First People’s Hospital of Yulin in Guangxi, China. We collected demographic characteristics, medical histories and biochemical indicators,including PA screening measurements (PAC, PRC, and ARR), renal function (serum creatinine, blood urea nitrogen, uric acid), and electrolytes (serum potassium, serum sodium, serum chloride, serum calcium). eGFR was calculated as 175 × (SCR) −1.234 × (Age)−0.179 (×0.79 if female) using the modified MDRD equation tailored for the Chinese population ^[18]^. We excluded the participants with renal diseases, including renal cancer, obstructive nephropathy, nephritis, nephrotic syndrome, polycystic kidney disease, nephrectomyand other urological disorders, as these conditions could alter eGFR levels. Additionally, pariticipants who were pregnant, suffering from severe cardiovascular or cerebrovascular disease, malignancy, and usage of mineralocorticoid receptor antagonists were further excluded. Finally, our study included a total of 612 participants based on 1:1 propensity score matching by age and gender, each of PA group, EH group, and NC group comprised of 204 participants, respectively.

### Ascertainment of Primary Aldosteronism

According to guidelines^[19]^, before conducting the initial PA screening, participants with hypokalemia needed to remedy the levels of serum potassium exceedto 3.5 mmol/L. Patients who taking antihypertensive medications, such as dihydropyridine calcium channel blockers, β-blockers, Angiotensin-Converting Enzyme Inhibitors and Angiotensin II Receptor Blockers, were advised to switch to non-dihydropyridine or α-blockers medicines to control blood pressure for three weeks. In this study, ARR≥ 30 is considered as positive in the preliminary screening. Patients with positive screening resutlts were recommended to take the following confirmatory testing, 1) Intravenous saline loading test, when the PAC levels exceed to 100 pg/ml after the test, or the PAC suppression ratio is ≤30% was defined as positive test. 2)Captopril challenge test, when the PAC exceeds to 110 pg/ml at two hours post-test, or the PAC suppression ratio is ≤30% was defined as positive test. Two senior endocrinologist confirmed the final diagnosis of PA.

### Ascertainment of Essential Hypertension

EH is diagnosed based on the absence of identifiable secondary causes. The diagnostic process encompasses a comprehensive medical history and physical examination, supplemented by routine laboratory tests to exclude secondary factors like renal disease, adrenal gland disorders, and thyroid dysfunction. Moreover, the diagnosis involves systematically ruling out established causes of secondary hypertension, such as renal hypertension, renal artery stenosis, pheochromocytoma, Cushing’s syndrome, hyperthyroidism, and polyarteritis nodosa. Additionally, patients exhibiting transient increases in blood pressure, such as those with white coat hypertension, are excluded from this diagnosis to ensure accuracy.

### Mendelian Randomization Analysis

To investigate the causal relationship between PA and eGFR, bidirectional and multivariable two-sample MR analyses were conducted using summary-level data from genome-wide association studies (GWAS). Initially, univariable MR analysis was conducted in the forward and reverse directions between PA and eGFR to establish their causal relationships. Then, multivariable MR (MVMR) analysis, which incorporating of systolic blood pressure (SBP) and diastolic blood pressure (DBP), was further performed to assess the role of blood pressure in the causal relationship between PA and eGFR.

To our knowledge, only one publicly available GWAS dataset for PA exists in European populations, which comprising of 562 PA cases and 950 controls (GWAS catalog accession code GCST90129615)^[20]^. Genetic instrumental variables (IVs) for eGFR were derived from a large-scale GWAS study among over one million individuals (GCST008059)^[21]^. The GWAS summary statistics for DBP and SBP involving a total of 349,328 participants were obtained from GWAS datasets GCST90301698 and GCST90301695, respectively^[22]^. To avoid the MR results might be bia from racial heterogeneity, all participants used in these MR studieswere restricted to European descents.

For the summary GWAS statistics of eGFR, SBP and DBP, we selected genetic IVs with a significant *P* threshold of <5×10^-8^. For PA, we chose single nucleotide polymorphisms (SNPs) with a P value of <5×10^-6^ as IVs due to the scarcity of SNPs. Subsequently, IVs were clumped within a 10 Mb window using a stringent linkage disequilibrium (LD) threshold of r^2^ = 0.001 to ensure that SNPs were independent. Next, F statistics were employed to assess the strength of the genetically determined IVs, when F>10 to align with the first Mendelian Randomization assumption and to avoid bias from weak IVs^[23-25]^. Finally, we harmonized the data to ensure the effect of a SNP on the exposure and the effect of that SNP on the outcome correspond to the same allele. Detailed information on the IVs is provided in supplemental data.

We identified Inverse variance weighted (IVW) as primary analytical method^[26]^, and applied other MR methods, includinge Weighted Median (WM), to verify the robustness of the causal effects^[27]^. MR-Egger and MR-pleiotropy tests are utilized to assess horizontal pleiotropy ^[28]^,MR-PRESSO is applied to identify and remove outlier SNPs^[29]^ and MR-heterogeneity methods are utilized to detect population heterogeneity^[30]^.

### Statistical Analysis

To mitigate skewness and enhance statistical power, non-normally distributed variables such as aldosterone, renin, ARR, and eGFR were log-transformed. Differences between groups for skewed variables were assessed using the Mann-Whitney U test, while one-way ANOVA was utilized for normally distributed variables. Chi-square tests were applied for comparisons between categorical groups. Multiple linear regression was used to evaluate the differences in eGFR between groups,and stratified analysis of the PA group,adjusting covariates contained gender, age, BMI, serum potassium, and type 2 diabetes. Forest plots were used to visualize the effect sizes of the models. All statistical analyses were performed using R software (version 4.2.2), with the TwoSampleMR (version 0.5.6) and MR-PRESSO (version 1.0) packages for MR analyses. A two-sided P value of < 0.05 was considered statistically significant.

## RESULT

### Baseline Characteristics of the Participants

Of the 612 participants, the average age was 53.85 years old, and the proportions of females and those with T2DM were 66% and 57%, respectively. In the PA group, the median PAC was 185.09 pg/ml, the median PRC was 2.72 pg/ml, and the median ARR was 73.14. There were no significant differences among the three groups in blood urea nitrogen, uric acid, serum chloride, or serum calcium levels. However, eGFR, SBP, DBP, serum creatinine, BMI, serum potassium, serum sodium, and type 2 diabetes showed statistically significant differences among the groups. Notably, the SBP, DBP, and eGFR of the PA group were similar to those of the EH group but significantly different from the NC group(Table 1).

**Table 1.**
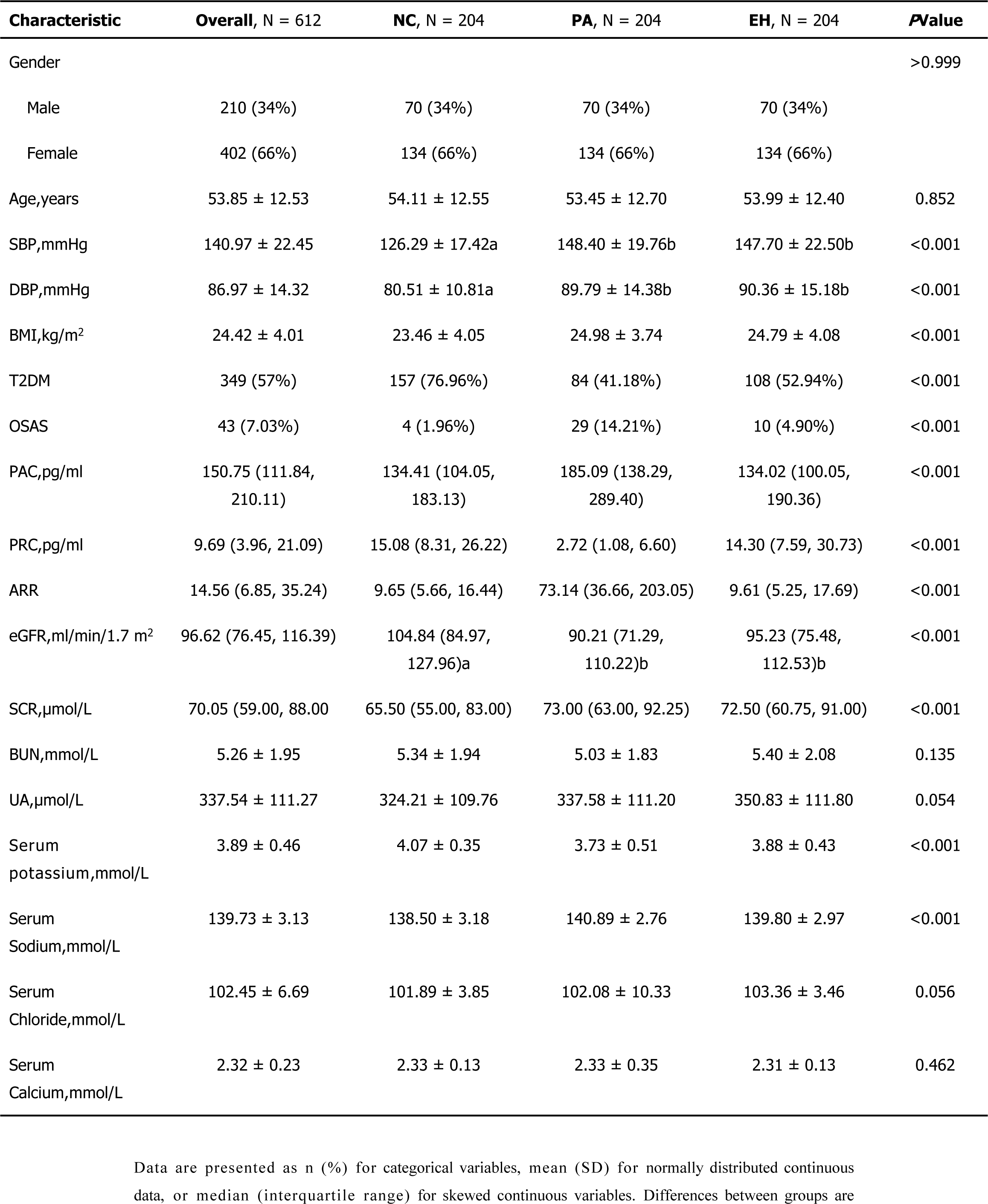
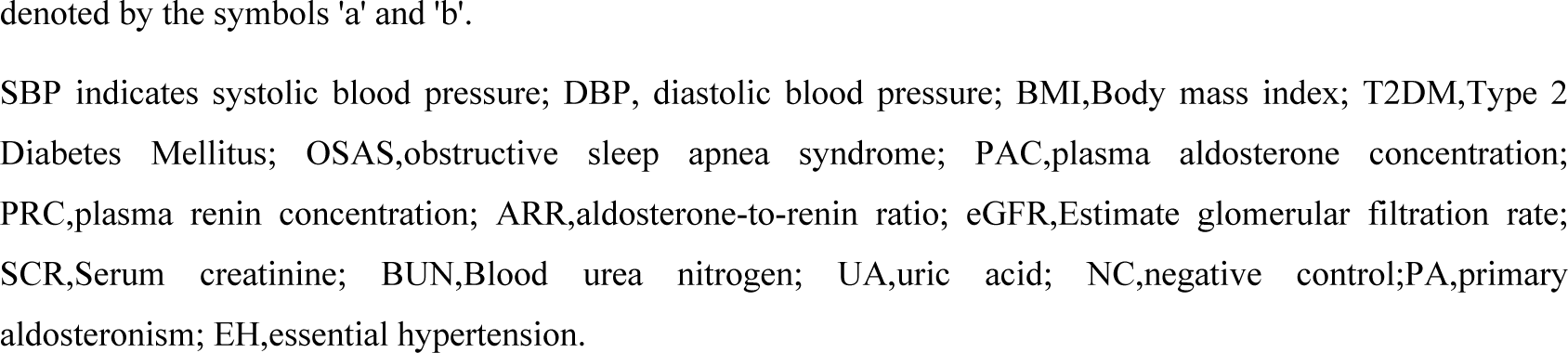
Baseline Characteristics of the Three Groups of Participants.

| Characteristic | Overall, N = 612 | NC, N = 204 | PA, N = 204 | EH, N = 204 | PValue |
| --- | --- | --- | --- | --- | --- |
| Gender |  |  |  |  | >0.999 |
| Male | 210 (34%) | 70 (34%) | 70 (34%) | 70 (34%) |  |
| Female | 402 (66%) | 134 (66%) | 134 (66%) | 134 (66%) |  |
| Age,years | 53.85 ± 12.53 | 54.11 ± 12.55 | 53.45 ± 12.70 | 53.99 ± 12.40 | 0.852 |
| SBP,mmHg | 140.97 ± 22.45 | 126.29 ± 17.42a | 148.40 ± 19.76b | 147.70 ± 22.50b | <0.001 |
| DBP,mmHg | 86.97 ± 14.32 | 80.51 ± 10.81a | 89.79 ± 14.38b | 90.36 ± 15.18b | <0.001 |
| BMI,kg/m<sup>2</sup> | 24.42 ± 4.01 | 23.46 ± 4.05 | 24.98 ± 3.74 | 24.79 ± 4.08 | <0.001 |
| T2DM | 349 (57%) | 157 (76.96%) | 84 (41.18%) | 108 (52.94%) | <0.001 |
| OSAS | 43 (7.03%) | 4 (1.96%) | 29 (14.21%) | 10 (4.90%) | <0.001 |
| PAC,pg/ml | 150.75 (111.84, 210.11) | 134.41 (104.05, 183.13) | 185.09 (138.29, 289.40) | 134.02 (100.05, 190.36) | <0.001 |
| PRC,pg/ml | 9.69 (3.96, 21.09) | 15.08 (8.31, 26.22) | 2.72 (1.08, 6.60) | 14.30 (7.59, 30.73) | <0.001 |
| ARR | 14.56 (6.85, 35.24) | 9.65 (5.66, 16.44) | 73.14 (36.66, 203.05) | 9.61 (5.25, 17.69) | <0.001 |
| eGFR,ml/min/1.7 m<sup>2</sup> | 96.62 (76.45, 116.39) | 104.84 (84.97, 127.96)a | 90.21 (71.29, 110.22)b | 95.23 (75.48, 112.53)b | <0.001 |
| SCR,μmol/L | 70.05 (59.00, 88.00) | 65.50 (55.00, 83.00) | 73.00 (63.00, 92.25) | 72.50 (60.75, 91.00) | <0.001 |
| BUN,mmol/L | 5.26 ± 1.95 | 5.34 ± 1.94 | 5.03 ± 1.83 | 5.40 ± 2.08 | 0.135 |
| UA,μmol/L | 337.54 ± 111.27 | 324.21 ± 109.76 | 337.58 ± 111.20 | 350.83 ± 111.80 | 0.054 |
| Serum potassium,mmol/L | 3.89 ± 0.46 | 4.07 ± 0.35 | 3.73 ± 0.51 | 3.88 ± 0.43 | <0.001 |
| Serum Sodium,mmol/L | 139.73 ± 3.13 | 138.50 ± 3.18 | 140.89 ± 2.76 | 139.80 ± 2.97 | <0.001 |
| Serum Chloride,mmol/L | 102.45 ± 6.69 | 101.89 ± 3.85 | 102.08 ± 10.33 | 103.36 ± 3.46 | 0.056 |
| Serum Calcium,mmol/L | 2.32 ± 0.23 | 2.33 ± 0.13 | 2.33 ± 0.35 | 2.31 ± 0.13 | 0.462 |

denoted by the symbols 'a' and 'b'.
SBP indicates systolic blood pressure; DBP, diastolic blood pressure; BMI,Body mass index; T2DM,Type 2 Diabetes Mellitus; OSAS,obstructive sleep apnea syndrome; PAC,plasma aldosterone concentration; PRC,plasma renin concentration; ARR,aldosterone-to-renin ratio; eGFR,Estimate glomerular filtration rate; SCR,Serum creatinine; BUN,Blood urea nitrogen; UA,uric acid; NC,negative control;PA,primary aldosteronism; EH,essential hypertension.

### Associations of PA and eGFR compared to control participants

Compared to the NC group, multiple linear regression model revealed that PA was associated with a decline eGFR (β = -0.234, [95% CI, -0.099, -0.039], P<0.001) after adjusted potential confounders, and the association remained significant even adjusted for SBP (β = -0.234, [95% CI,-0.102,-0.035], P<0.001) or DBP (β = -0.217, [95% CI,-0.095,-0.032], P<0.001). We further stratified the PA patients into three groups according to the levels of PAC, PRC and ARR to explore the correlations of PA related phenotypes with eGFR. When compared to the NC group, significant inverse associations were consistently observed between PAC,PRC and ARR with eGFR across three groups, except for the lowest tertile of PAC adjusted for blood pressure (P > 0.05) (Table 2).

**Table 2.** Associations of primary aldosteronism and estimated glomerular filtration rate compared to negative control group.

| Beta represents the change in eGFR(ml/min/1.7 m<sup>2</sup>) |  |  |  |  |  |  |  |  |
| --- | --- | --- | --- | --- | --- | --- | --- | --- |
|  | Model 1 |  | Model 2 |  | Model 3 |  | Model 4 |  |
|  | Beta (95% CI) | P Value | Beta (95% CI) | P Value | Beta (95% CI) | P Value | Beta (95% CI) | P Value |
| NC | Reference | - | Reference | - | Reference | - | Reference | - |
| PA | - 0.248 ( -0.1 , -0.045 ) | <0.001 | - 0.234 ( -0.099 , -0.039 ) | <0.001 | -0.234(-0.102,-0.035) | <0.001 | -0.217(-0.095,-0.032) | <0.001 |
| PA Subgroups* |  |  |  |  |  |  |  |  |
| PAC T1 | -0.161(-0.091,-0.014) | 0.008 | -0.134(-0.083,-0.004) | 0.029 | -0.123(-0.083,0.003) | 0.066 | -0.124(-0.081,0) | 0.05 |
| PAC T2 | -0.219(-0.11,-0.033) | <0.001 | -0.228(-0.114,-0.036) | <0.001 | -0.275(-0.134,-0.047) | <0.001 | -0.252(-0.124,-0.042) | <0.001 |
| PAC T3 | -0.262(-0.134,-0.052) | <0.001 | -0.278(-0.15,-0.051) | <0.001 | -0.311(-0.165,-0.059) | <0.001 | -0.274(-0.15,-0.047) | <0.001 |
| PRC T1 | -0.303(-0.148,-0.067) | <0.001 | -0.25(-0.133,-0.048) | <0.001 | -0.272(-0.145,-0.052) | <0.001 | -0.261(-0.139,-0.05) | <0.001 |
| PRC T2 | -0.156(-0.089,-0.012) | 0.01 | -0.204(-0.105,-0.026) | 0.001 | -0.245(-0.124,-0.035) | 0.001 | -0.203(-0.107,-0.024) | 0.002 |
| PRC T3 | -0.18(-0.098,-0.02) | 0.003 | -0.162(-0.098,-0.009) | 0.018 | -0.171(-0.104,-0.009) | 0.02 | -0.159(-0.098,-0.007) | 0.024 |
| ARR T1 | -0.166(-0.093,-0.016) | 0.006 | -0.168(-0.098,-0.013) | 0.01 | -0.184(-0.107,-0.014) | 0.011 | -0.166(-0.099,-0.011) | 0.015 |
| ARR T2 | -0.155(-0.088,-0.012) | 0.01 | -0.19(-0.1,-0.023) | <0.001 | -0.204(-0.109,-0.024) | 0.002 | -0.187(-0.101,-0.021) | 0.003 |
| ARR T3 | -0.317(-0.152,-0.072) | <0.001 | -0.273(-0.145,-0.053) | <0.001 | -0.308(-0.162,-0.062) | <0.001 | -0.284(-0.151,-0.055) | <0.001 |

\*Each row under the
PA subgroups represents a different subgroup of PA patients according to the tertiles of PAC, PRC, and ARR.
Model 1: no adjustment. Model 2: adjusted for gender, age, BMI, type 2 diabetes mellitus, and serum potassium. Model 3: adjusted for systolic blood pressure based on Model 2. Model 4: adjusted for diastolic blood pressure based on Model 2. eGFR, Estimate glomerular filtration rate; PAC, plasma aldosterone concentration; PRC, plasma renin concentration; ARR, aldosterone-to-renin ratio; NC, negative control; PA, primary aldosteronism.

### Associations of PA and eGFR compared to patients with EH

Compared to the EH group, there were no significant difference between PA and eGFR (β = -0.065, [95% CI,-0.046,0.009], P=0.217). When stratified the PA patients into three groups according to the levels of PAC, PRC and ARR, patients in the highest PAC groups (β = -0.146 [95% CI, -0.093, -0.008], p=0.021), the lowest PRC group (β = -0.127 [95% CI, -0.084, -0.004], P=0.033), and the highest ARR group (β = -0.147 [95% CI, -0.092, -0.009], 0.017) had lower eGFR compared to the EH group, and these associations remained significant even further adjusted for SBP or DBP (Table 3).

**Table 3.**
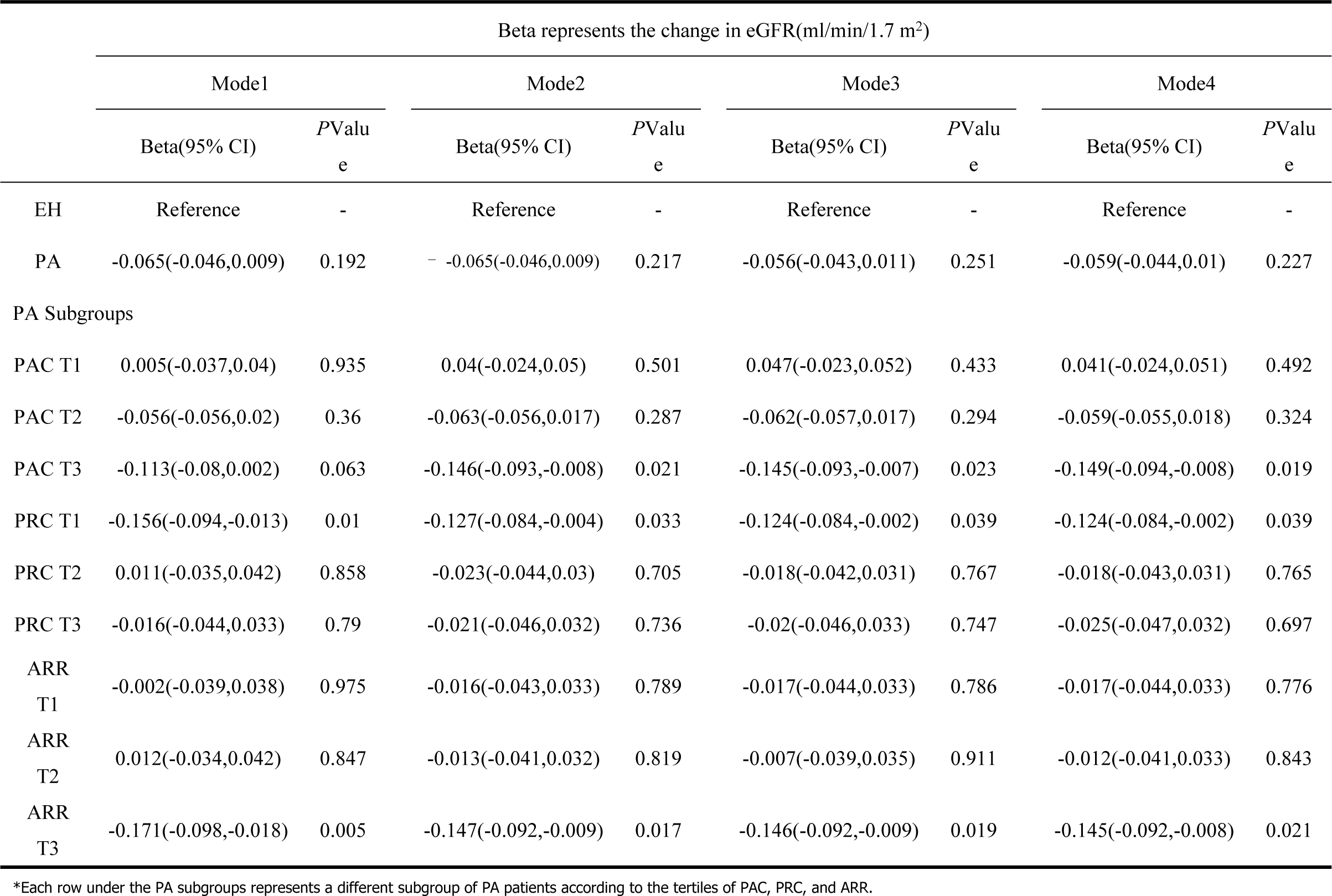
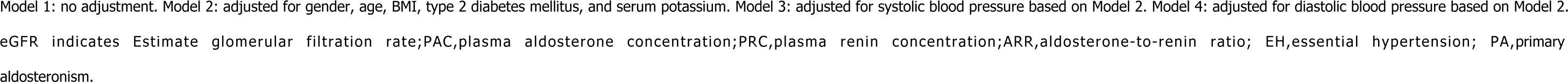
Associations of primary aldosteronism and estimated glomerular filtration rate compared to essential hypertension group.

| Beta represents the change in eGFR(ml/min/1.7 m<sup>2</sup>) |  |  |  |  |  |  |  |  |
| --- | --- | --- | --- | --- | --- | --- | --- | --- |
|  | Mode1 |  | Mode2 |  | Mode3 |  | Mode4 |  |
|  | Beta(95% CI) | PValue | Beta(95% CI) | PValue | Beta(95% CI) | PValue | Beta(95% CI) | PValue |
| EH | Reference | - | Reference | - | Reference | - | Reference | - |
| PA | -0.065(-0.046,0.009) | 0.192 | -0.065(-0.046,0.009) | 0.217 | -0.056(-0.043,0.011) | 0.251 | -0.059(-0.044,0.01) | 0.227 |
| PA Subgroups |  |  |  |  |  |  |  |  |
| PAC T1 | 0.005(-0.037,0.04) | 0.935 | 0.04(-0.024,0.05) | 0.501 | 0.047(-0.023,0.052) | 0.433 | 0.041(-0.024,0.051) | 0.492 |
| PAC T2 | -0.056(-0.056,0.02) | 0.36 | -0.063(-0.056,0.017) | 0.287 | -0.062(-0.057,0.017) | 0.294 | -0.059(-0.055,0.018) | 0.324 |
| PAC T3 | -0.113(-0.08,0.002) | 0.063 | -0.146(-0.093,-0.008) | 0.021 | -0.145(-0.093,-0.007) | 0.023 | -0.149(-0.094,-0.008) | 0.019 |
| PRC T1 | -0.156(-0.094,-0.013) | 0.01 | -0.127(-0.084,-0.004) | 0.033 | -0.124(-0.084,-0.002) | 0.039 | -0.124(-0.084,-0.002) | 0.039 |
| PRC T2 | 0.011(-0.035,0.042) | 0.858 | -0.023(-0.044,0.03) | 0.705 | -0.018(-0.042,0.031) | 0.767 | -0.018(-0.043,0.031) | 0.765 |
| PRC T3 | -0.016(-0.044,0.033) | 0.79 | -0.021(-0.046,0.032) | 0.736 | -0.02(-0.046,0.033) | 0.747 | -0.025(-0.047,0.032) | 0.697 |
| ARR T1 | -0.002(-0.039,0.038) | 0.975 | -0.016(-0.043,0.033) | 0.789 | -0.017(-0.044,0.033) | 0.786 | -0.017(-0.044,0.033) | 0.776 |
| ARR T2 | 0.012(-0.034,0.042) | 0.847 | -0.013(-0.041,0.032) | 0.819 | -0.007(-0.039,0.035) | 0.911 | -0.012(-0.041,0.033) | 0.843 |
| ARR T3 | -0.171(-0.098,-0.018) | 0.005 | -0.147(-0.092,-0.009) | 0.017 | -0.146(-0.092,-0.009) | 0.019 | -0.145(-0.092,-0.008) | 0.021 |

\*Each row under the PA subgroups represents a different subgroup of PA patients according to the tertiles of PAC, PRC, and ARR.
Model 1: no adjustment. Model 2: adjusted for gender, age, BMI, type 2 diabetes mellitus, and serum potassium. Model 3: adjusted for systolic blood pressure based on Model 2. Model 4: adjusted for diastolic blood pressure based on Model 2. eGFR indicates Estimate glomerular filtration rate; PAC, plasma aldosterone concentration; PRC, plasma renin concentration; ARR, aldosterone-to-renin ratio; EH, essential hypertension; PA, primary aldosteronism.

### Subgroup Analysis of PA related phenotypes and eGFR in PA patients

Among PA patients, multiple linear regression showed that, after adjusted potential confounders,PAC (β = -0.137, [95% CI,-0.209,-0.065], P<0.001) and ARR (β = -0.03, [95% CI,-0.061,0], P=0.048) were negatively associated with eGFR. Compared to the lowest tertile group, patients in the highest tertile of PAC was associated with lower eGFR (T3 vs. T1, β = -0.061, [95% CI,-0.108,-0.015], P=0.01), and the trend was statistically significant (P-trend = 0.013). In subgroup analyses, we found that PA patients who were male (β =-0.069, [95% CI,-0.109,-0.028], P=0.001) and aged 60 and older(β = -0.081, [95% CI,-0.123,-0.04], P<0.001) resulted in lower eGFR. (Table 4 and Figures 1)

**Figure 1:**
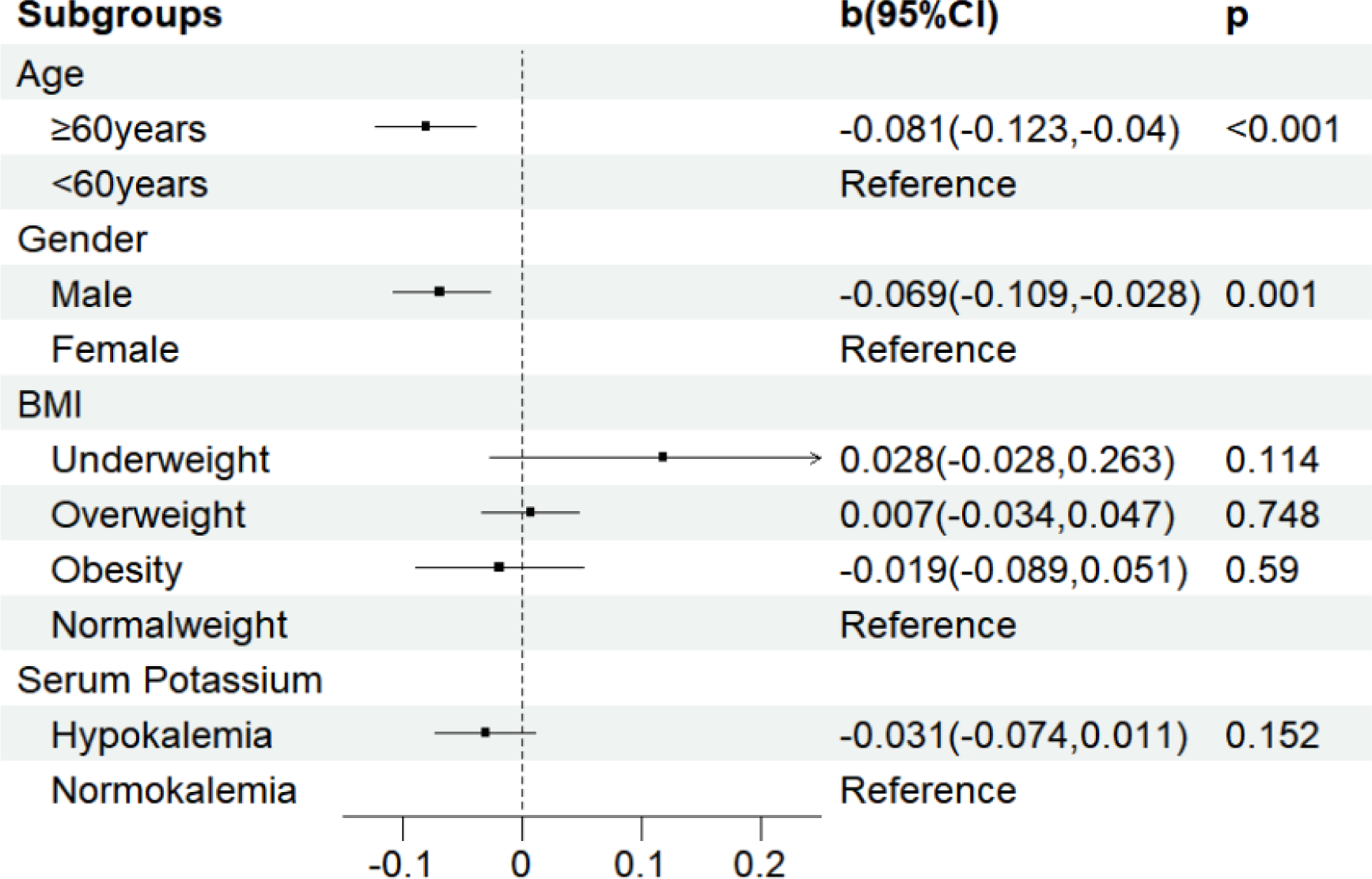
Subgroup analysis of the association between PA and eGFR among PA patients,adjusted for covariates. Adjusted covariates included gender、age、type 2 diabetes mellitus and BMI. BMI(kg/m^2^) indicates Body mass index.underweight:BMI<18.5;normalweight:18.5 ≤ BMI<25;overweight:25 ≤ BMI<30;obesity:BMI ≥ 30; hypokalemia:serum potassium level below 3.5 mmol/L.

**Table 4.**
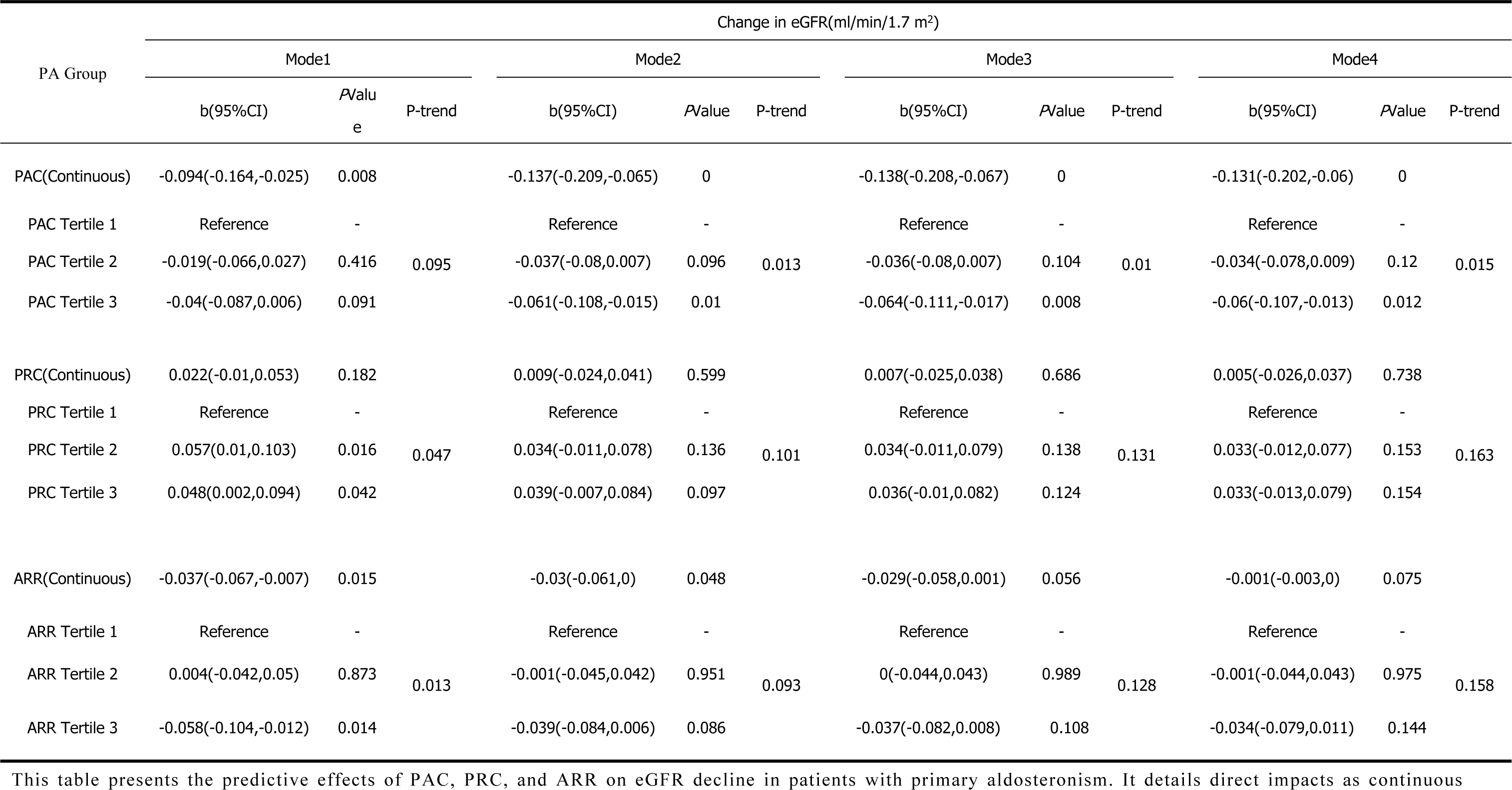
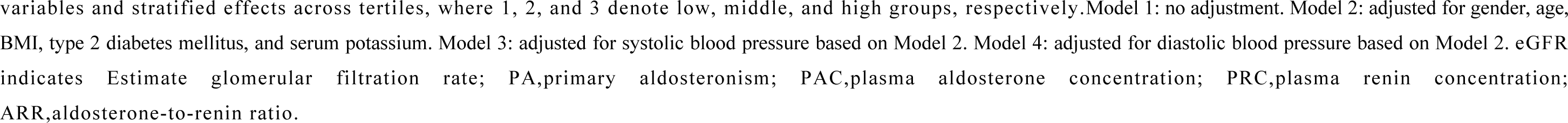
Correlations of ARR, PRC, and PAC with Estimated Glomerular Filtration Rate (eGFR) in Primary Aldosteronism.

| PA Group | Change in eGFR(ml/min/1.7 m<sup>2</sup>) |  |  |  |  |  |  |  |  |  |  |  |
| --- | --- | --- | --- | --- | --- | --- | --- | --- | --- | --- | --- | --- |
|  | Mode1 |  |  | Mode2 |  |  | Mode3 |  |  | Mode4 |  |  |
|  | b(95%CI) | PValue | P-trend | b(95%CI) | PValue | P-trend | b(95%CI) | PValue | P-trend | b(95%CI) | PValue | P-trend |
| PAC(Continuous) | -0.094(-0.164,-0.025) | 0.008 |  | -0.137(-0.209,-0.065) | 0 |  | -0.138(-0.208,-0.067) | 0 |  | -0.131(-0.202,-0.06) | 0 |  |
| PAC Tertile 1 | Reference | - |  | Reference | - |  | Reference | - |  | Reference | - |  |
| PAC Tertile 2 | -0.019(-0.066,0.027) | 0.416 | 0.095 | -0.037(-0.08,0.007) | 0.096 | 0.013 | -0.036(-0.08,0.007) | 0.104 | 0.01 | -0.034(-0.078,0.009) | 0.12 | 0.015 |
| PAC Tertile 3 | -0.04(-0.087,0.006) | 0.091 |  | -0.061(-0.108,-0.015) | 0.01 |  | -0.064(-0.111,-0.017) | 0.008 |  | -0.06(-0.107,-0.013) | 0.012 |  |
| PRC(Continuous) | 0.022(-0.01,0.053) | 0.182 |  | 0.009(-0.024,0.041) | 0.599 |  | 0.007(-0.025,0.038) | 0.686 |  | 0.005(-0.026,0.037) | 0.738 |  |
| PRC Tertile 1 | Reference | - |  | Reference | - |  | Reference | - |  | Reference | - |  |
| PRC Tertile 2 | 0.057(0.01,0.103) | 0.016 | 0.047 | 0.034(-0.011,0.078) | 0.136 | 0.101 | 0.034(-0.011,0.079) | 0.138 | 0.131 | 0.033(-0.012,0.077) | 0.153 | 0.163 |
| PRC Tertile 3 | 0.048(0.002,0.094) | 0.042 |  | 0.039(-0.007,0.084) | 0.097 |  | 0.036(-0.01,0.082) | 0.124 |  | 0.033(-0.013,0.079) | 0.154 |  |
| ARR(Continuous) | -0.037(-0.067,-0.007) | 0.015 |  | -0.03(-0.061,0) | 0.048 |  | -0.029(-0.058,0.001) | 0.056 |  | -0.001(-0.003,0) | 0.075 |  |
| ARR Tertile 1 | Reference | - |  | Reference | - |  | Reference | - |  | Reference | - |  |
| ARR Tertile 2 | 0.004(-0.042,0.05) | 0.873 | 0.013 | -0.001(-0.045,0.042) | 0.951 | 0.093 | 0(-0.044,0.043) | 0.989 | 0.128 | -0.001(-0.044,0.043) | 0.975 | 0.158 |
| ARR Tertile 3 | -0.058(-0.104,-0.012) | 0.014 |  | -0.039(-0.084,0.006) | 0.086 |  | -0.037(-0.082,0.008) | 0.108 |  | -0.034(-0.079,0.011) | 0.144 |  |

This table presents the predictive effects of PAC, PRC, and ARR on eGFR decline in patients with primary aldosteronism. It details direct impacts as continuous
variables and stratified effects across tertiles, where 1, 2, and 3 denote low, middle, and high groups, respectively. Model 1: no adjustment. Model 2: adjusted for gender, age, BMI, type 2 diabetes mellitus, and serum potassium. Model 3: adjusted for systolic blood pressure based on Model 2. Model 4: adjusted for diastolic blood pressure based on Model 2. eGFR indicates Estimate glomerular filtration rate; PA, primary aldosteronism; PAC, plasma aldosterone concentration; PRC, plasma renin concentration; ARR, aldosterone-to-renin ratio.

### Two-Sample MR and MVMR Analysis of PA and eGFR

In the forward MR analysis, IVW method indicated that genetically predicted higher risk of PA was causally associated with declined eGFR (β=-6.67×10^-4^;[95%CI, -1.29×10^-3^ to -4.33×10^-5^], P = 0.036); other MR analyses. such as WMM, further corroborated the direction of their causal relationship (Figure 2). Besides, Cochran’s test and MR-Egger regression showed no evidence of heterogeneity (Q = 4.936, P = 0.668) or directional pleiotropy (intercept = -0.0009, P = 0.331). In the reverse MR analysis, IVW method revealed that genetically predicted eGFR did not have an impact on PA (β = -1.093, [95% CI,-10.946,8.778], P =0.828) (Figure 2), the robustness of the MR findings were further supported by sensitivity tests.

**Figure 2.**
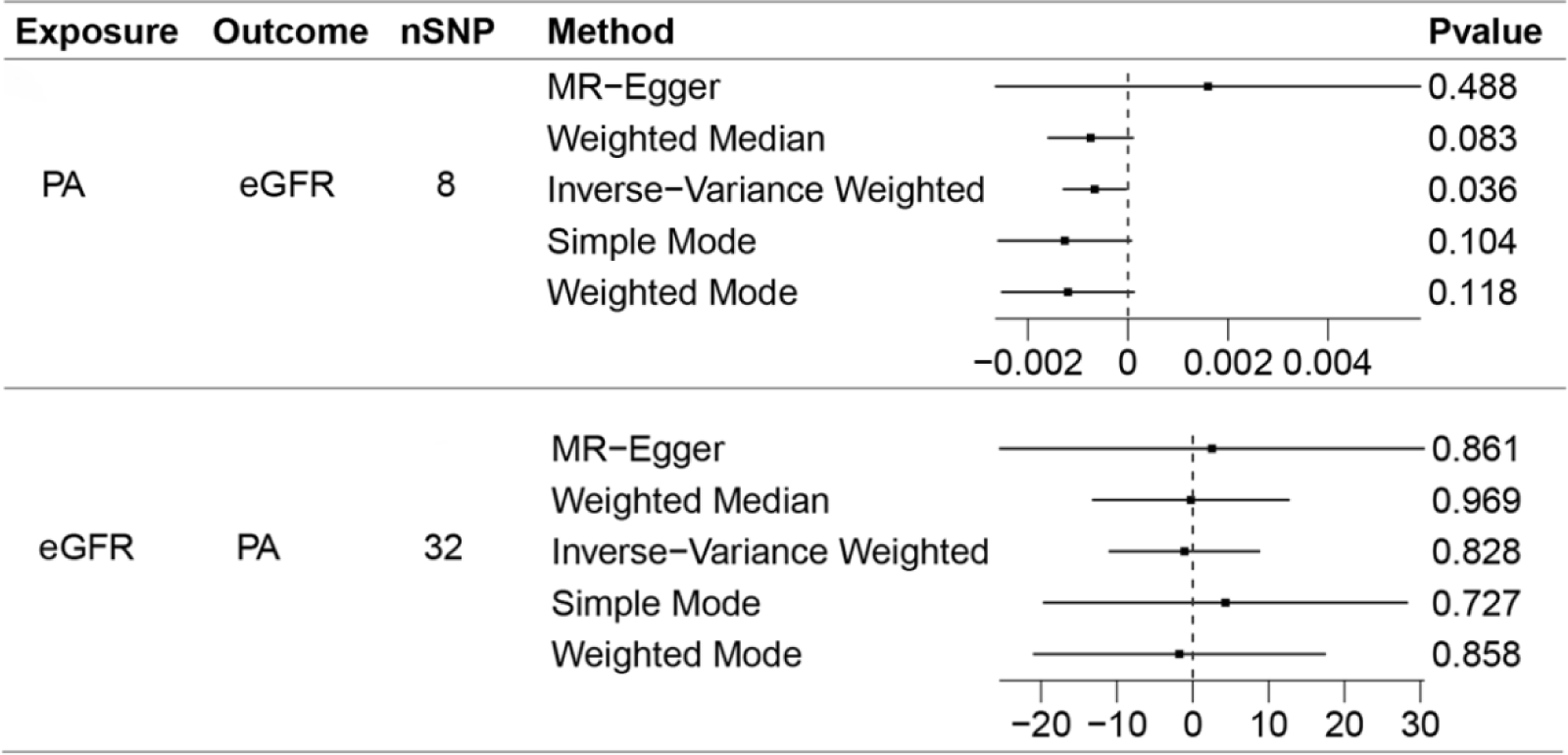
Bidirectional two-sample MR analyses of genetically predicted PA and eGFR. PA indicates primary aldosteronismle;eGFR,estimated glomerular filtration rate.

Subsequently, MVMR analysis was further conducted to explore the roles of SBP and DBP in the effect of genetic susceptibility to PA on eGFR (Figure 3). IVW method showed that genetically predicted PA remained a causal relationship with a declined eGFR (β = -1.12×10^-3^;[95% CI, -2.13×10^-3^ to -1.11×10^-4^]; P = 0.029) after adjusting for SBP. The association of PA and eGFR turn into null after adjusting for DBP (β = -6.67×10^-4^; P = 0.494), but the significance recovered statistically when selecting PA-associated SNPs with a P value of = 0.005 as the IVs (β = -1.54×10^-3^; 95%CI,[-2.69×10^-3^to-3.91×10^-4^],P=0.008). No significant heterogeneity or horizontal pleiotropy was observed in the MVMR analysis (P>0.05).

**Figure 3.**
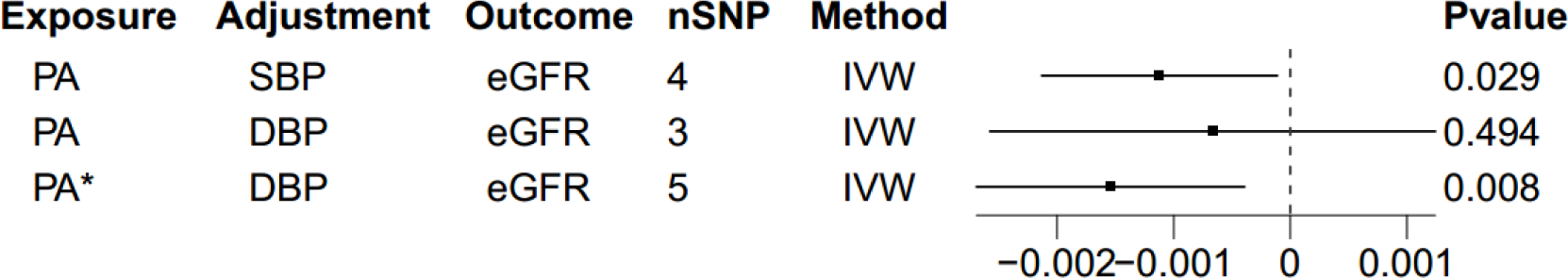
Multivariable two-sample MR analysis of genetically predicted PA and eGFR. The analysis adjusts for two scenarios: SBP or DBP. The notation “PA*” indicates that genetic instrumental variables for PA were selected based on a significance threshold of P=0.005. PA indicates primary aldosteronis;eGFR,estimated glomerular filtration rate.SBP,Systolic Blood Pressure;DBP,Diastolic Blood Pressure;IVW,inverse-variance weighted.

## DISCUSSION

In this study, the cross-sectional data firstly demonstrated that patients with PA resulted in much lower eGFR than those of age- and sex-matched negative controls or EH patients, especially in the extremely severe condition of PA that characterized by the highest levels of PAC and ARR, or the lowest levels of PRC. Of note, these inverse associations remained significant even adjusted for blood pressure. Moreover, bidirectional two-sample MR analyses confirmed that genetically predicted higher risk of PA was causally associated with eGFR decline, and MVMR analysis further clarified that the impacts of PA on eGFR might not mediated by blood pressure. These findings collectively suggest that PA plays a crucial role in the reduction of eGFR, and these adverse effects might be independent of blood pressure.

Recent evidence has been reported that patients with PA was associated with more severe renal impairment and a higher risk of chronic kidney disease than those with EH^[31-34]^,few studies focus on the relationship between PA and continuous variable eGFR, however. Our cross-sectional findings indicated that eGFR was significantly lower in PA patients compared to the matched negative controls or EH patients, especially in the PA patients with extremely severe condition. Consistent with our findings, Reincke et al found that eGFR was significantly lower in PA patients than matched EH patients in a case-control study, a finding echoed in recent analyses examining PA-related kidney damage^[7,35]^. Catena et al. observed that the positive association between low renin levels and lower eGFR in PA patients^[36]^, suggests that the severity of PA is associated with lower eGFR, which further validates our findings. Conversely, Ribstein et al. found no significant difference in GFR between tumor-related PA and EH patients with similar demographic data and duration of hypertension^[13]^. The contradictory findings regarding PA and eGFR could be attribute to several reasons. Firstly, accumulating evidence has recognized PA as a progressive disease that target organ damage intensifies with the severity of PA rather than a binary state^[37,38]^. Secondly, relative increases in PAC leading to sodium retention may cause an initial elevation in eGFR due to hyperfiltration, maintaining normal metabolic functions at milder stages of PA, However, interventions that mitigate this hyperfiltration may reveal the true extent of PA-related renal impairment^[14,39]^. When the severity of PA surpasses the kidney’s compensatory abilities, a decline in eGFR occurs. Epidemiological studies are often subject to confounding factors, leading to inconsistent conclusions.MR minimizes potential biases due to confounding factors and reverse causation, thereby increasing the reliability of the results^[17]^. Therefore, we conducted a two-sample MR analysis to further elucidate the relationship between PA and eGFR. The IVW results indicate a causal association between PA and declined eGFR, while eGFR is not associated with PA. Sensitivity analyses confirmed the robustness of these findings.

It is worth noting that is that the trend of eGFR decline due to PA remained signigicant even adjusting for SBP or DBP compared to the normotension controls and EH groups. Furthermore, MVMR analysis also verified that SBP could not alter the causal association between genetically predicted PA and decreased eGFR, and a similar pattern of effect was detected in DBP adjusted model when we relaxed the threshold of the risk variants associated with PA to include more SNPs as IVs. These findings revealed that PA could affect the reduction of eGFR independent of blood pressure pathway. Clinical studies have found that PA led to renal glomerular damage and progressive glomerulosclerosis, interstitial inflammation and scarring, ultimately worsenrenal function^[40,41]^; In fundamental research, aldosterone cause renal impairment through mechanisms such as promoting inflammation^[42]^, oxidative stress^[43]^, fibrosis^[44]^, mesangial cell proliferation^[45]^, and podocyte damage^[46]^. Therefore, we believed that the underlying mechanisms involving in PA-driven eGFR decline might not blood pressure but the inflammatory response and fibrosis caused by autonomous aldosterone secretion. Besides, we found only PAC was significantly correlates with the reduction of eGFR among PA patients, which could corroborate with the above explanations to some extent.

In subgroup analysis, we found that PA patients who were males or older than 60 years old have a greater likelihood of eGFR decline. In fact, eGFR typically decreases with increasing age in healthy populations^[10]^, and renal function becomes more impaired after suffering from PA. Related studies have observed that male PA patients are more susceptible to the reduction in eGFR^[47]^, which could be potentially attributate to endocrine regulation. Animal studies suggested that testosterone may accelerate the progression of renal disease, whereas estrogen appears to offer renal protection, manifested in its anti-inflammatory and anti-fibrotic effects^[48]^ and its role in regulating glomerular endothelial cell functions^[49]^,these factors may account for the diminished influence of PA on females relative to males,However, the mechanisms underlying these gender differences in PA patients require further investigation to elucidate.

Our study has the following strengths. Firstly, a well-designed case-control study, which included sex- and age-matched NC controls, patients with EH and PA, is helpful to understand the actual effects of PA on eGFR decline independent of blood pressure. Furthermore, to our best knowledge, this is the first study to systematically assess the causal relationship between PA and eGFR through bidirectional two-sample MR analysis, and MVMR analysis was further performed to elucidate the mediated effects of blood pressure in the causal relationship between PA and eGFR. These analyses make the MR results more credibility and robustness. However, our study also has several limitations. Firstly, epidemiological data are derived from the Chinese population, while the MR analysis is based on European populations, indicating potential population heterogeneity. Therefore, it is necessary to conduct a PA GWAS study specifically in the Chinese population.Secondly, we did not document the specific dosages of antihypertensive medications for all participants at enrollment, making the assessment of the direct effects of these medications on eGFR is difficult. Lastly, owing to our study is a cross-sectional study with small sample size, long-term follow-up in larger population is necessary to further clarify the long-term effects of PA on eGFR, However, our study does provide some evidence for the association between PA and eGFR.

In summary, our study highlighted that PA is causally associated with lower eGFR independent of blood pressure, and the adverse effects might be greater than negative controls or EH patients.In the treatment of PA, it may be necessary to pay more attention to the control of PAC, particular attention should be directed towards the protection of renal function in older or male patients with PA. Future efforts to establish a high-quality PA GWAS dataset are crucial for elucidating the pathogenesis of PA and its associations with other diseases.

## Ethics statement

This study was received ethical approval by the Ethics and Human Subject Committee of Guangxi Medical University, China (No. 2022-0193), and the Ethics and Human Subject Committee of First Affiliated Hospital of Guangxi Medical University, China (No. 2023-K090-01). All participants provided written informed consents at the time of their recruitment.

## Data Availability

Data cannot be shared publicly because of privacy concerns. Data are available from the Ethics Committee of Guangxi Medical University (contact via Zengnan Mo) for researchers who meet the criteria for access to confidential data.

## Acknowledgments

The study was performed in the first Affiliated Hospital of Guangxi Medical University and the First People’s Hospital of Yulin in Guangxi, China. We sincerely thank all the study participants and the staff involved in facilitating and conducting the study.

